# Electrocardiographic Digital Biomarkers in Asymptomatic Schoolchildren with Rheumatic Heart Disease

**DOI:** 10.64898/2026.02.16.26346419

**Authors:** Amsalu Tomas Chuma, Jens-Uwe Voigt, Ahmed S. Youssef, Melkamu H. Asmare, Chunzhuo Wang, Carolina Varon, Desalew M. Kassie, Rik Willems, Bart Vanrumste

## Abstract

Subclinical rheumatic valvular disease is a significant yet underdiagnosed contributor to the global rheumatic heart disease (RHD) burden. Early detection through population screening is essential to prevent its progression to severe RHD. Rhythm changes and prolongations of PR and QTc intervals in the ECG are described in the advanced RHD cases. However, these parameters were not yet studied in asymptomatic RHD. We aimed to investigate the potential of ECG biomarkers for screening RHD in asymptomatic schoolchildren. ECG tracings from 611 schoolchildren aged 10 to 20 years were selected from a cohort screened for RHD in four schools in an RHD-endemic region. Confirmatory diagnoses were based on echocardiographic findings, where 564 (F=326, M=238) were healthy, and 47 (F=28, M=19) were positive for RHD (24 borderline RHD and 23 definite RHD). Independent, blinded reviewers manually annotated the ECGs and PR interval (PR), P-wave dispersion (PWd), and the ratio between the P-wave duration and PR interval (Pw/PR) were analyzed. The mean age of the study cohort at diagnosis was 16.1 ± 2.5 years, and 58% of the participants were females. Atrial fibrillation was seen in 8% (n=4), and prolonged PR in 2% (n=1) of RHD-positive cases. The mean ± std for normals vs RHD is (PR, 138±19 vs 150±19), (Pw/PR, 0.75±0.06 vs 0.71±0.07), and (PWd, 49±14 vs 56±17). The PR (p<0.001), Pw/PR (p<0.001), and PWd (p=0.008) showed a significant difference between healthy and RHD-positive subjects. The PR was increased consistently with severity across age groups above and below 16 years. The PR, PWd, and Pw/PR can serve as non-invasive biomarkers for the screening of RHD in at-risk schoolchildren. Monitoring alterations in these markers at an early stage of RHD is crucial for enabling prompt management and follow-up. It is thus evident that ECG can support an intermittent ambulatory RHD screening in resource-limited settings.

## 1 Introduction

Rheumatic heart disease (RHD) remains a major global health concern in low- and middle-income countries, leading to significant morbidity and mortality among pediatric populations. Echocardiographic evaluation is the gold standard for diagnosis. Despite treatment advances, the lack of effective preventive measures, inadequate healthcare infrastructure, and delayed diagnosis in resource-limited settings exacerbate the disease burden. A substantial proportion of the disease burden arises from subclinical cases that progress silently without clinical symptoms. A study in Uganda reported that subclinical RHD was detected in 21.5% of children screened by echocardiography, compared to 0.8% diagnosed with clinical RHD [1]. Similar studies in [2],[3] also showed that subclinical RHD was four times more common than clinically recognized RHD in school-aged children. Specific manifestations occurring in Acute Rheumatic Fever (ARF), such as carditis, prolonged PR interval, and Sydenham chorea, are strongly associated with an elevated risk of progression to RHD [4], [5]. While these features highlight the importance of periodic screening programs to detect early RHD cases in at-risk populations, preventive policies in many low-resource settings are hindered by a lack of disease awareness, limited access to echocardiography, and trained personnel to interpret results [6]. Consequently, the potential use of electrocardiography (ECG) as a screening tool for RHD in such settings may be both beneficial and challenging. The challenge is because of the complex relationship between electrical disturbances and non-specific ECG parameter alterations that overlap with other cardiac or systemic conditions, thereby complicating the accurate identification of RHD-specific patterns [7], [8].

The significance of ECG parameters (see Figure 1 panel (C)) and their changes in RHD, ARF, and rheumatic carditis have been well established in the literature. Common ECG findings include prolonged PR interval, T-wave inversion, QT interval prolongation, T-wave alternans (TWA), prolonged TpTe interval, and TpTe/QT ratio, which reflect varying degrees of myocardial and pericardial inflammation. These electrical disturbances often correlate with the active inflammatory phase of the disease and may serve as early indicators of subclinical cardiac involvement. For instance, studies have shown that prolongation of PR, QT, and the T-wave inversions were common in patients with rheumatic fever. These changes were often transient, corresponding with the active inflammatory phase of the disease. However, the abnormalities persisted in some cases, suggesting the presence of ongoing myocardial involvement. In patients with ARF and RHD, prolonged PR intervals also persisted [7], [8], indicating varying degrees of atrioventricular (AV) conduction delay in the disease’s later stages. The severity of repolarization abnormalities, including QT and TpTe prolongation, has also been shown to correlate with the extent of myocardial inflammation, reinforcing the diagnostic potential of ECG markers in the detection of RHD [9], [10].

**Figure 1.**
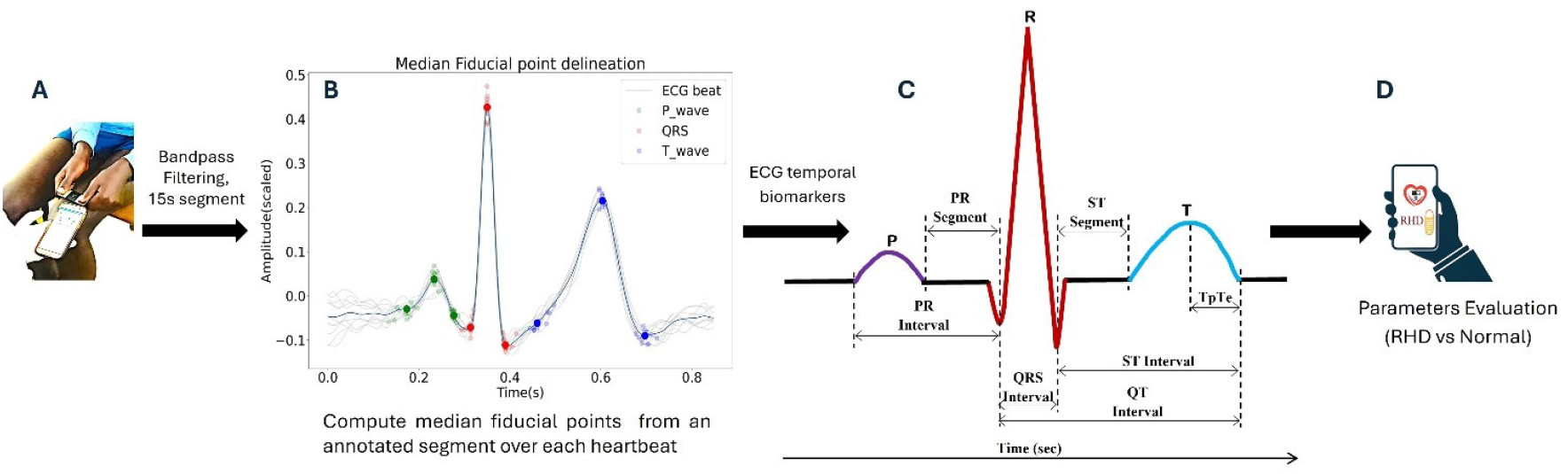
Study workflow. Panel (A): ECG signal acquisition using a KM sensor connected via Bluetooth to a smartphone. Panel (B): Annotated PQRST fiducial points for each beat (grey lines) and averaged waveform centered on R-peaks (black line); median fiducial points shown as bold circles. Panel (C): Measured ECG intervals and segments. Panel (D): Comparison of ECG parameters between normal and RHD-positive subjects.

Furthermore, rheumatic mitral stenosis or aortic regurgitation can cause significant hemodynamic stress, which results in chronic left atrial enlargement or left ventricular volume overload. Such structural heart changes may predispose patients to atrial and ventricular arrhythmias. Although the arrhythmias commonly associated with RHD are atrial fibrillation or other supraventricular arrhythmias [5], severe valve dysfunction could alter ventricular repolarization, possibly creating a substrate for ventricular arrhythmias. Robyns, T., et al. [11] reported that higher levels of the index of cardio-electrophysiological balance (iCEB) were seen in conditions that raise the risk of Torsades de Pointes (TdP). Conversely, lower iCEB levels are more frequently linked with the development of ventricular tachycardia (VT) or ventricular fibrillation (VF) that are not mediated by TdP. The study [11] demonstrated that the index of cardio-electrophysiological balance (iCEB) levels increase under conditions that predispose individuals to Torsades de Pointes (TdP), whereas a decrease in iCEB is more likely to result in non-TdP mediated ventricular tachycardia (VT) or ventricular fibrillation (VF). Their findings suggest the importance of a balanced relationship between depolarization and repolarization to maintain ventricular electrical stability.

P-wave dispersion (PWd) is the difference between the longest and shortest P-wave durations. PWd is associated with uneven sinus impulse conduction and increased AF risk [12]. This marker reflects atrial inhomogeneity, which can be caused by left atrial dilation, fibrosis, and structural disruptions from mitral valve disease or rheumatic inflammation, resulting in prolonged P-wave duration and dispersion on ECG [13], [14]. Studies show a positive correlation between maximum P-wave duration and left atrial size [15], though its role as a predictor of AF is debated. Conversely, the minimum P-wave duration is negatively correlated with mitral valve area in patients with rheumatic mitral stenosis (MS) [15], [16]. This lower limit has been reported to be similar in patients with MS and healthy subjects, indicating that atrial electromechanical delay is associated with left atrial size rather than severity of MS. In the early stages of RHD, structural remodeling of the atria is uncommon [5], [17], so prolonged or bifid P-waves are not prominent enough to be detected. Nevertheless, inflammation of the myocardium during recurrent attacks causes AV conduction delay [6]–[8], [18]. As a result, PWd and the ratio of atrial conduction time to AV conduction time provide insight that combines the effects of early and acute RHD.

The presence of TWA has also been reported as a non-invasive marker of arrhythmogenic potential in rheumatic myocarditis [19]. The findings from [20] indicated significantly prolonged TpTe intervals and TpTe/QT ratios in children with acute rheumatic carditis compared with healthy controls. Analogously, these parameters were also significantly prolonged in patients with mitral valve prolapse, indicating disturbances observed in the early stages of RHD [21]. Rheumatic mitral regurgitation and stenosis are widely recognized as significant contributors to cardiac electrical instability in patients with RHD. The relationship between RHD precursor diseases such as rheumatic fever, acute rheumatic carditis, and ventricular arrhythmogenesis is complex and multifactorial. Assessing the significance of ECG markers in at-risk children with subclinical RHD supports preventive mass screening programs and cost-effective clinical management.

Portable ECG devices such as KardiaMobile 6L ECG sensor (AliveCore Inc., Mountain View, California, USA), hereafter referred to as KM, are affordable in RHD prone regions, and parameter changes may serve as a valuable non-invasive marker for detecting subclinical RHD. The KM sensor has demonstrated high concordance with the standard 12-lead ECG in detecting specific cardiac parameters, particularly in adults with atrial fibrillation (AF). Studies by [22]–[24] reported a detection agreement of 99% for AF, making KM a reliable tool for arrhythmia monitoring. Similarly, the KM ECG sensor showed strong performance in identifying congestive heart failure through PR interval measurements (90%) and QRS dispersion, with a 90% agreement. The corrected QT interval (QTc) showed slight variability, but within a clinically acceptable range of 20 ms, with 98% accuracy.

Similarly, smartwatches such as the Apple Watch with single-lead ECG have been validated for AF detection. For instance, Pepplinkhuizen S., et al. [25] reported a sensitivity of 93.5% and specificity of 100% in patients scheduled for cardioversion, compared with the 12-lead ECG gold standard. Another comparative study [26] of three smartwatch ECGs (Apple Watch Series 5®, Samsung Galaxy Watch Active 3®, and Withings Move ECG®) found that the Samsung Galaxy Watch Active 3 achieved 88% sensitivity and 81% specificity for AF when analyzed by automated software (versus a 12-lead ECG). These findings suggest that low-cost ECGs are effective tools for detecting rhythm disturbances and variations in interval dispersion. Despite this potential, their utility remains constrained by occasional inconclusive recordings and limited diagnostic accuracy for detection of structural abnormalities. A major advantage of such portable ECG devices lies in their affordability and scalability, which enable widespread population-level screening for conditions such as ARF, thereby facilitating early preventive interventions. Nonetheless, changes in key ECG intervals and dispersion parameters have not been systematically investigated in the early stages of RHD, leaving a critical gap in understanding their diagnostic value for detecting subclinical disease.

This study included pediatric patients aged 10-20 years from an ambulatory population-based RHD screening in a high-prevalence region of Ethiopia. Demographic data, clinical characteristics, and measurements were collected using the KM sensor. We analyzed three key ECG parameters: PR interval duration, P-wave duration (PWd), and the P-wave-to-PR interval ratio (Pw/PR). Because the KM sensor records single-lead ECG data, we refer to variations in these measurements as temporal variability rather than true dispersion, which requires multiple leads. The primary outcome was RHD status, determined by echocardiography according to the 2012 World Heart Federation criteria [17] (see Appendix A). Participants were classified as either normal (healthy) or RHD positive, with the latter including both borderline and definite RHD cases. The study aims to investigate whether these ECG parameters differ between RHD-negative and RHD-positive children, assess their associations with disease severity, and evaluate their potential as screening markers for detecting subclinical RHD in asymptomatic children.

## 2 Materials and Methods

### 2.1 Dataset

A total of 611 schoolchildren and young adults aged 10 to 20 years (mean ± SD: 16.1 ± 2.4 years) from rural areas of Ethiopia were randomly selected and screened for RHD. Each participant underwent electrocardiography (ECG), cardiac auscultation, and echocardiography for confirmatory diagnosis. The echocardiographic recordings were independently reviewed by two cardiologists, and diagnostic classification was performed according to the 2012 echocardiography-based guidelines for RHD diagnosis [17]. Participants with a history of any known congenital or acquired cardiac disease, previous cardiac surgery, or systemic illness affecting cardiac function were excluded. Only participants meeting these criteria and providing written informed consent or parental consent for minors were included in the study. Based on the diagnostic assessments, the final cohort comprised 47 RHD-positive participants and 564 RHD-negative or healthy participants. The healthy group consisted of those with normal echocardiographic findings. Among the RHD-positive cases (F = 28; M = 19), 51.1% (n=24) were classified as borderline RHD and 48.9% (n=23) as definite RHD. The analysis of electrocardiographic parameters within this cohort is outlined in Section II-B.

### 2.2 Workflow

The overall workflow diagram is illustrated in Figure 1. First, ECG recordings for each participant were obtained using a commercial CE-marked KM sensor (see panel (A)). The KM sensor has two leads at the front and a reference at the back. Each recording has a 30-second duration, and a 15-second waveform from lead II of the KM was used for evaluation. After assessing the qualities of ECGs using the Neurokit algorithm [27], the P, QRS, and T waves (see panel (B)) were manually annotated by two blinded reviewers for subsequent analysis, as described in Section II-E. The annotations consisted of the P, QRS, and T waves onset, peak, and offset fiducial points.

### 2.3 Ethical considerations

This study was approved by the Ethical Committee of UZ Leuven (REGNR: B3222022001075). Local permissions were granted by Soddo Christian Hospital in Ethiopia (REF: SCH/194/10/15), other government administrative offices, and the participating schools. All data collection procedures were conducted in accordance with the General Data Protection Regulation (GDPR, EU 2016/679), and appropriate approvals were obtained prior to initiating the study. Written informed consent was obtained from parents or guardians, and assent was obtained from all participating students.

### 2.4 Statistical analysis

Statistical analyses were conducted using R software (version 4.1.0) [28]. Continuous variables were summarized as means ± standard deviations for normally distributed data and medians for non-normally distributed data. Normality was assessed using the Shapiro-Wilk test. For comparison of continuous variables between two groups, the Welch’s t-test was employed for normally distributed data, and the Mann-Whitney U test for non-normally distributed data. Categorical variables were presented as counts and percentages. Correlations between continuous variables were assessed using Pearson’s correlation for normally distributed data and Spearman’s rank correlation for non-normally distributed data. Analysis of variance (ANOVA) was used to compare percentages across the Normal, borderline, and definite RHD groups. The association between ECG parameters and outcomes was evaluated using different multivariate models. The p-values on two-tailed alpha < 0.05 were considered statistically significant.

### 2.5 ECG temporal parameters

Two independent, blinded annotators manually labeled at least 10 beats from each of the 15-second ECG tracings in Lead II of the KM sensor. Manual annotation of P, QRS, and T wave fiducial points is inherently subjective. To mitigate this, discrepancies between annotators at each fiducial point, such as mismatched markings, were resolved by calculating the mean of the available annotations to establish a consensus. Furthermore, to enhance consistency across multiple beats, the median values of these annotations were employed. ECG measurements of key parameters, shown in Figure 1 panel(C), were subsequently computed from these consensus fiducial points for further analysis. It is important to note that PWd values from a single-lead may show limited temporal variability in P-wave duration that loosely reflects atrial conduction stability. Hence, dispersions computed here represent intra-record beat-to-beat variability.

The PR interval was measured as the distance between the onset of the P-wave and the onset of the QRS-wave. The QT interval was determined as the distance between the onset of the QRSwave and the offset of the T-wave. The QTc was calculated using Bazett’s formula [29]. In cases where there is no horizontal segment between the end of the T wave and the beginning of the P-wave due to a rapid heartbeat, a horizontal line through the PQ segment is used as a reference for the isoelectric line to begin a reasonable QRS onset and offset [30], [31]. However, some recordings have a rising downward PQ segment and no clear RS-T junctions to determine the onsets and offsets of the QRS-wave. Therefore, the transition between the rapidly descending and ascending terminal limb of the QRS was used to maintain consistency across subjects. The P-wave is measured according to the recommendations in [12] from the point of the first detectable upward or downward slope from the isoelectric line for positive or negative waveforms, respectively, to the return point to the isoelectric line.

Prolongation of PR is determined by age and heart rate adjusted references [32]. The Pw/PR is measured as the ratio of P-wave to PR duration. The PWd was calculated as the difference between the maximum and minimum values of the P-wave durations. Because relying on a single maximum or minimum value may be disproportionately influenced by outliers, PWd was alternatively estimated by calculating the difference between the mean of the two shortest P-wave durations and the mean of the two longest P-wave durations. This approach provides a more robust assessment of atrial conduction variability by mitigating the impact of extreme values.

## 3 Results

The baseline characteristics of the study population are summarized in Table I. The mean age at the time of data acquisition was 16.1 ± 2.5 years, with females comprised 58% (n = 354) of the study participants. The age distribution was skewed towards older adolescents, with approximately 15% of participants aged 14 years or younger. There was no statistically significant difference in BMI or mean heart rate between the RHD-positive and healthy groups. After establishing cohort characteristics, waveform annotation was assessed as shown in Figure 1. Incidence of rhythm abnormalities were seen in 8%(n=4) of RHD-positive cases. In addition, prolonged PR was detected in only 2% (n=1) of RHD positives.

**Table I:** Characteristics of the study participants between RHD positive and healthy subjects (Mean ± SD)

| Variables | Healthy (Mean ± SD)<br>(n=564) | RHD positives (Mean ± SD)<br>(n=47) |
| --- | --- | --- |
| Age (years) | 16.1 ± 2.5 | 16.7 ± 2.5 |
| 10-13 | 14.4% | 0.7% |
| 14-16 | 42.3% | 95.3% |
| 17-20 | 43.3% | 4.0% |
| Sex |  |  |
| Female | 326 | 28 |
| Male | 238 | 19 |
| BMI (kg/m<sup>2</sup>) | 19.5 ± 2.7 | 19.3 ± 2.7 |
| Mean HR (bpm) | 78 ± 14 | 79 ± 13 |

Data distribution and statistics for the assessed key ECG parameters in both groups are shown in Table II. There was no strong association between the parameters and sex or disease outcome, and no significant difference in QTc interval with age. However, QTc requires gender-specific thresholds because normative QTc values differ by sex. Thus, the QTc differed by gender, with females having a longer QTc than males (445ms ± 25ms vs 423ms ± 28ms; p <0.001). Box plots for both PwRHD and normal groups are illustrated in Figure 2.

**Table II:** Comparison of temporal ECG parameters in RHD positive and healthy groups (Mean ± SD) in ms.

| Variables | Normal (ms)<br>(n=564) | RHD-positive (ms)<br>(n=47) | p-value |
| --- | --- | --- | --- |
| QTc |  |  |  |
| Female | 445 ± 26 | 445 ± 18 | 0.668 |
| Male | 422 ± 28 | 419 ± 26 | 0.671 |
| Pw | 102 ± 13 | 105 ± 12 | 0.159 |
| QRSd | 93 ± 15 | 92 ± 16 | 0.911 |
| Tw | 196 ± 23 | 200 ± 24 | 0.234 |
| PR | 138 ± 19 | 150 ± 19 | **<0.001** |
| PWd | 49 ± 19 | 56 ± 17 | **0.008** |
| Pw/PR | 0.75 ± 0.06 | 0.71 ± 0.07 | **<0.001** |
| TpTe/QTc | 0.21 ± 0.02 | 0.20 ± 0.03 | **0.031** |
| QTcd | 4.28 ± 2.9 | 4.83 ± 3.1 | 0.326 |

Pw : P-wave duration, Tw : T wave duration. QTcd: QTc dispersion. The Pw/PR and TpTe/QTc values have no units. Statistically significant p-values are bolded.

**Figure 2.**
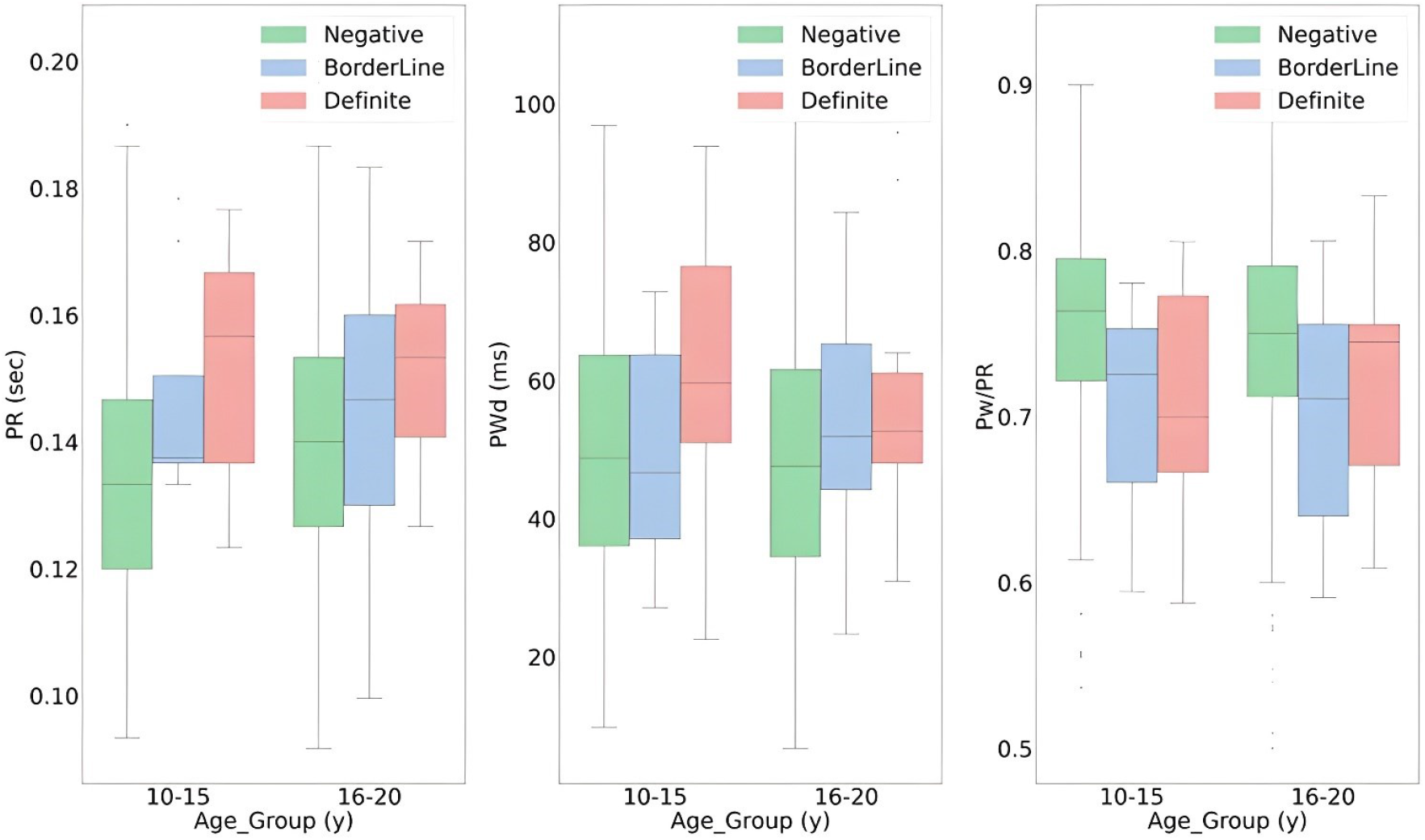
RHD severity per age groups (≤15 and>15y) for PR (p<0.001), PWd (p=0.008) and Pw/PR (p<0.001) ratio parameters. The box-and-whisker plots illustrate the distribution of PR, PWd, and PW/PR among participants classified as Normal, Borderline, and Definite RHD for two age-groups (≤15 and *>*15y). The central line within each box denotes the median, the box boundaries represent the IQR, and the whiskers extend to 1.5×IQR from the lower and upper quartiles. Outliers beyond this range are plotted as individual dots.

Participants in the RHD-positive group had prolonged PR (138ms ± 19ms vs 150ms ± 19ms), increased PWd (49 ± 19 vs 56 ± 17), and a lower Pw/PR ratio (0.75 ± 0.06 vs 0.71 ± 0.07). An increased trend of values for PR was observed in disease outcomes. Both PR (p<0.001) and PWd (p=0.008) indicate strong evidence of differences in depolarization and intra- and interatrial conduction times between the RHD-positive and healthy groups. Similarly, the Pw/PR also showed significant evidence (p<0.001) that RHD-positive subjects had a lower ratio than those who had normal echocardiographic findings. In contrast, the QTc, QTc dispersions (QTcd), and wave durations such as P-wave (Pw), QRS complex (QRSd), and T-wave (Tw) showed no significant difference between normal and RHD positive groups as evaluated in Table II.

An ANOVA was performed to examine the effects of age group (10–15 years vs. 16–20 years) and disease severity (normal, borderline, and definite) on PR interval. The age groups were defined based on reported age-related differences in cardiac physiology and anatomy [32]. The ANOVA and post-hoc results for PR, PWd, and Pw/PR parameters were shown in Appendix B. For the PR interval, the overall model was statistically significant (F(5, 605) = 4.93, p<0.001) across six groups (severity×age-groups). There was a strong main effect of disease severity on PR (F(2, 605) = 8.39, p<0.001, η2 = 0.027), indicating that PR increased progressively with increasing disease severity. Post hoc comparisons showed that the borderline RHD group had a significantly higher PR than the normal healthy group (mean difference = 11ms, p = 0.012), and the definite RHD group also exhibited a higher PR compared to the normal group (mean difference = 17ms, p < 0.001).

The difference between borderline and definite groups was not statistically significant (mean difference = 6ms, p= 0.334), suggesting that conduction delay plateaus beyond the borderline stage. Similarly, ANOVA examining the effects of age group and severity revealed a statistically significant overall model for Pw/PR (F(5, 605) = 6.00, p<0.001), whereas no significant association was observed for PWd (F(5, 605) = 4.93, p<0.088). Post hoc analyses indicated that the definite group exhibited significantly higher PWd values compared with the normal group (mean difference = 13.88 ms, p = 0.010), as well as higher Pw/PR ratios (mean difference = 0.045, p = 0.018).

However, the difference between borderline and definite groups was not statistically significant for both PWd and Pw/PR parameters. The main effect of age group was not statistically significant (F(1, 605) = 0.058, p = 0.809, η2 ≈ 0.000), indicating that PR did not differ meaningfully between the 10– 15 and 16–20 year groups. Similarly, the interaction between disease severity and age group was not significant overall (F(2, 605) = 1.05, p = 0.352, η2 = 0.003), suggesting that the relationship between disease severity and PR was consistent across both age categories.

## 4 Discussion

The potential use of ECG parameters, including PR, PWd, and Pw/PR, for screening for RHD among at-risk schoolchildren was assessed in a randomly selected asymptomatic cohort. Our findings show that there was a significant difference in the mean for these parameters and in the TpTe/QTc ratio between those diagnosed with RHD and those with normal echocardiographic findings, whereas QTc did not differ significantly. The elevated TpTe/QTc ratio and increased PWd in RHD-positive cases suggest subclinical myocardial and atrial involvement. These results indicate that PR, Pwd, and Pw/PR could serve as potential digital biomarkers for early identification of children at higher risk of RHD during intermittent screening.

The PR interval is a sensitive marker of AV nodal conduction system involvement. Patients who have experienced ARF often present with rhythm and conduction abnormalities that may persist and evolve into RHD. In children with ARF, these abnormalities are pronounced and manifest in various ECG morphologies, including an elongated PR interval and AF [33], [34]. In line with the studies in [5]–[7], the statistical tests on PR, Pwd, and Pw/PR have shown strong evidence that there is a difference in mean values between healthy and RHD-positive participants. Nevertheless, only a limited number of positive cases, 8%, had rhythm disturbances and prolonged PR in the asymptomatic cohort.

Moreover, the findings in this study suggest that PWd and Pw/PR are also significant predictors of RHD positive outcome. The study in [13] reported that a PWd ≥ 40ms indicates heterogeneous atrial electrical activity, which is associated with an increased risk of atrial arrhythmias. Such complications are often present in patients with advanced and symptomatic RHD. A comparable study [15] demonstrated that patients with rheumatic MS exhibited a marginally higher mean PWd value of 55ms compared with healthy subjects, who demonstrated a mean PWd value of approximately 51ms. Our findings align with these claims, as the mean PWd in RHD-positive subjects was 56ms, which was statistically significantly higher than in normals (49ms).

On the other hand, an elevated QTc also reflects an increased cardiac electrical instability in patients with ventricular remodelling, fibrosis, and valvular dysfunction. The dispersion of QTc in patients with severe RHD has been reported to be higher [8], and [35] indicated that such elevated dispersion is associated with greater severity in patients with rheumatic mitral valve disease. Furthermore, elevated QTcd values in patients with ARF have been linked to an elevated risk of developing chronic RHD. However, our findings show no significant elevation of either QTc or QTcd in the RHD-positive group compared with healthy groups. A reason for this could be the subclinicality and early stages of the disease. Nevertheless, our analysis further confirms that TpTe/QTc is significantly different between RHD positives and healthy groups. This finding may reflect myocardial inflammation during acute rheumatic carditis, which leads to heterogeneous ventricular involvement and, consequently, increased repolarization dispersion, consistent with observations reported in [9], [10], [20].

It is noteworthy that no significant correlation was identified between age group and severity for any of the ECG parameters. These findings indicate that although ECG abnormalities are predictors of disease severity and prognosis, their impact does not notably diverge between younger (10–15 years) and older (16–20 years) individuals diagnosed with RHD. This finding contrasts with some previous research, which indicated that advanced age is associated with more severe RHD due to cumulative valve damage from recurrent ARF episodes. However, the absence of this interaction effect in this cohort may indicate a narrow age range and a relatively early disease stage within the pediatric population. The influence of gender on the etiology of RHD is still unclear. Nevertheless, observational evidence from multiple large-scale screening and registry studies indicates that clinically manifest RHD is more common in females than in males [36]. As shown in Table I, our findings indicate that females with RHD exhibit prolonged intervals and an elevated risk of RHD in comparison to their male counterparts with RHD.

Furthermore, as shown in the ANOVA results, no significant interaction between age groups and disease severity was observed. This indicates that age variations do not meaningfully influence the relationship between severity and the biomarkers PR, PWd, and Pw/PR within the study cohort. The observed differences in PR, PWd, and Pw/PR between healthy individuals and those with definite RHD reflect distinct pathophysiological alterations specific to established disease, whereas the absence of similar differences between borderline and definite RHD reflects the inability of the biomarkers to clearly distinguish the more subtle changes that occur in intermediate transition stages.

Overall, ECG biomarkers can be useful for rapid population-wide screening for RHD in resource-limited endemic regions. Nevertheless, its diagnostic sensitivity can be improved by applying machine learning algorithms for automated interpretation. The integration of both digital biomarkers from ECG, such as PR, PWd, and Pw/PR, into the established RHD standard of care has the potential to enhance disease prevention strategies and optimize patient management. Although echocardiography is the gold standard for diagnosing RHD, its use is limited by cost and accessibility in the Global South. Cost-effective tools such as ECG can serve as initial screening, complemented by confirmatory echocardiography to detect subclinical cases early.

## 5 Limitation

The findings of this study should be interpreted in light of the following limitations. First, a relatively small sample size for severe disease outcomes may have resulted in limited statistical power to detect all significant associations. Furthermore, the study was conducted in regions with high disease prevalence, characterized by overcrowded living conditions and lower socioeconomic status. These contextual factors may limit the applicability of the results in other unseen settings with improved healthcare infrastructure.

Second, observer bias is an inherent limitation of manual ECG annotation, particularly in delineating fiducial points such as P-waves, QRS complexes, and T-wave offsets. This subjectivity can compromise the accuracy of subsequent quantitative analyses and introduce inconsistencies in the training of machine learning models. While using median values of fiducial points helps mitigate the influence of outliers, it does not entirely eliminate subjectivity bias. Future work should aim to overcome this limitation by employing automated PQRST annotation methods. Despite these constraints and the limitations of the available data, the need for cost-effective screening tools to curb the widespread impact of RHD remains undeniable.

Third, because the KM device lacks precordial leads, parameters such as the Tpeak–Tend interval (TpTe) and the TpTe/QTc ratio, commonly measured from precordial leads, were derived from limb leads in this study. This methodological adjustment may limit the accuracy of repolarization assessment, as limb leads do not fully represent regional ventricular repolarization gradients captured in standard 12-lead ECGs. Similarly, the use of a single-lead for dispersion measures, such as PWd or QTcd, provides only a limited approximation of spatial electrical heterogeneity, which conventionally requires multiple lead vectors. Therefore, the reported repolarization and dispersion values should be interpreted cautiously, as they may differ from those obtained with conventional 12-lead measurements.

## 6 Conclusion

The findings of this study showed that the PR interval, PWd, and Pw/PR ratio are valuable non-invasive biomarkers for screening RHD in at-risk schoolchildren. These markers are significant independent predictors of positive outcomes of early RHD. The significance of assessing atrioventricular depolarization heterogeneity is useful for stratifying early RHD risk. Timely diagnosis, secondary prophylaxis, and continuous monitoring of subclinical cases should be prioritized in public health strategies to reduce the global burden of this preventable condition. Therefore, prolonged PR, elevated PWd, and decreased Pw/PR can be considered red flags for a poor prognosis, guiding local healthcare workers to adopt periodic screening for early disease detection in resource-limited settings.

## Data Availability

All data produced in the present study are available upon reasonable request to the authors.

## Acknowledgment

The study is funded by KU Leuven, center for affordable healthcare with funding reference REF23123123. The authors would like to thank the study participants, schools and concerned government bureaus. The authors are also grateful to the study team members and the collaborating hospitals University hospitals Leuven in Belgium, Tikur Anbesa Teaching Hospital and Soddo General Christian Hospital in Ethiopia, and Flanders AI for their unwavering support. We also want to note the use of generative AI tool for optimizing the manuscript. The dataset can be accessed to evaluate various models upon request.

## Author Contributions

Amsalu Tomas: Conceptualized the study, performed computer programming, collected dataset, conducted experiments and prepared the first draft manuscript. Melkamu Hunegnaw, Desalew Mekonnen, Carolina Varon, Rik Willems and Chunzhuo Wang: Frame dataset acquisition, analyze the experimental settings and results, and reviewed the manuscript. Jens-Uwe Voigt and Bart Vanrumste: Conceptualized the study, designed methodology, analyzed the experiments and results, and reviewed the manuscript.

## Ethics approval and consent to participate

The study was conducted according to the guidelines of the Declaration of Helsinki and approved by the Ethics Committee of the University hospital Leuven with reference number (B3222022001075) and the institutional review board of Soddo General Christian Hospital with reference number (SCH1941015). Clinical trial number is not applicable. Written informed consent was obtained from the patient(s) to publish the study results.

## Data Availability Statement

The dataset analyzed in this manuscript is not publicly available. However, it can be provided upon reasonable request via.

## APPENDIX

### A. 2012 WHF criteria for echocardiographic diagnosis of RHD, adapted from [17]

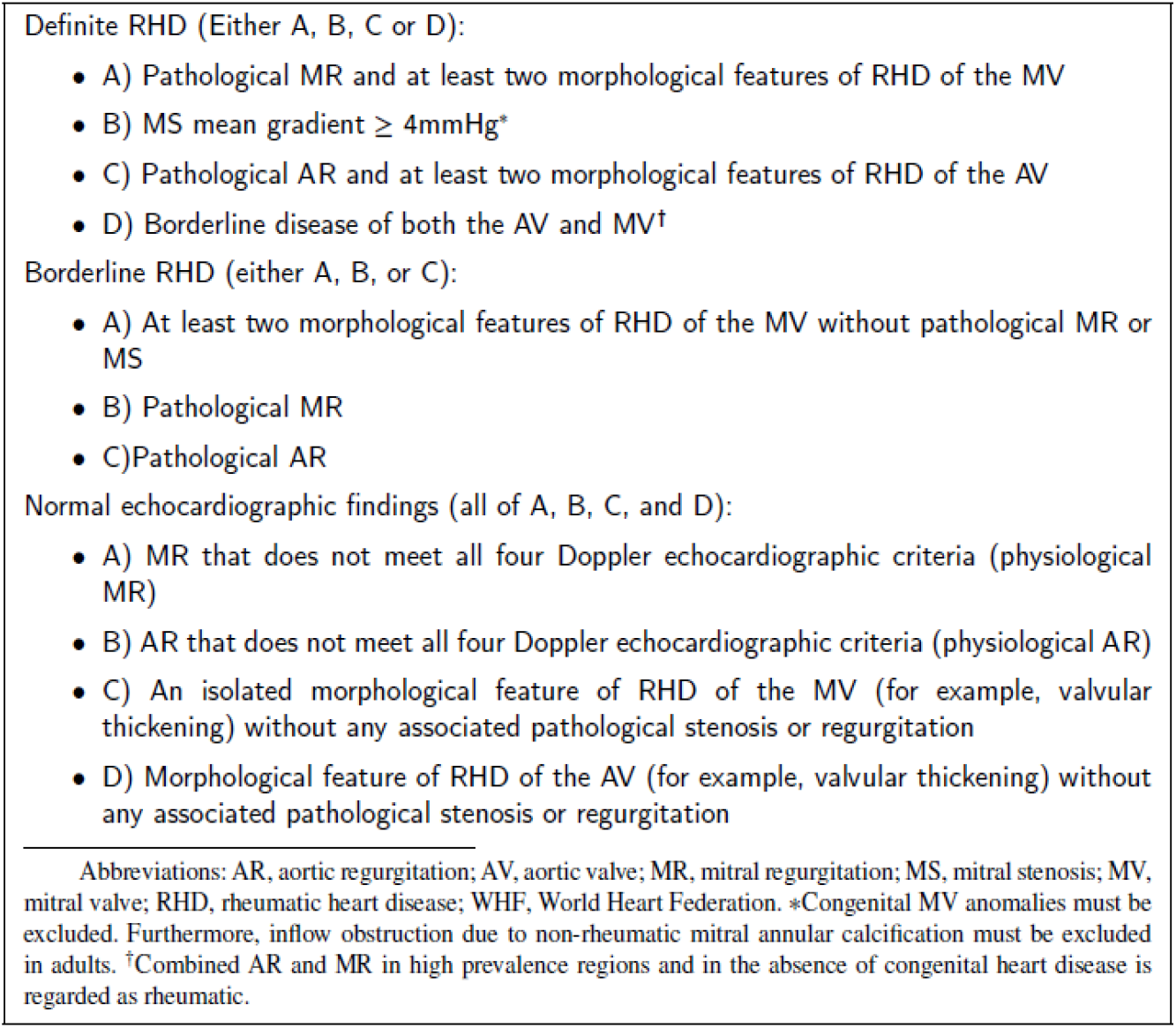

### B. ANOVA and Post-hoc results for PR parameter between disease severity and age groups (≤15y and >15y)

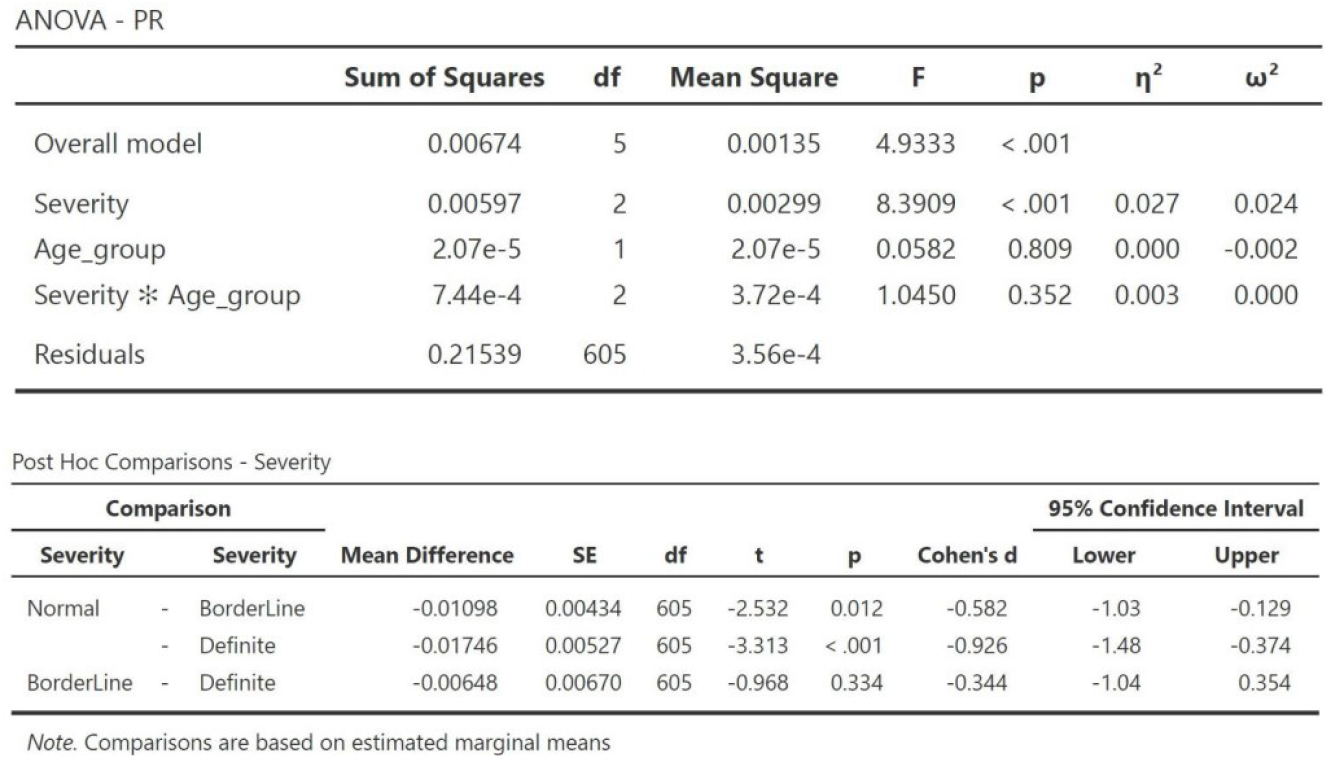
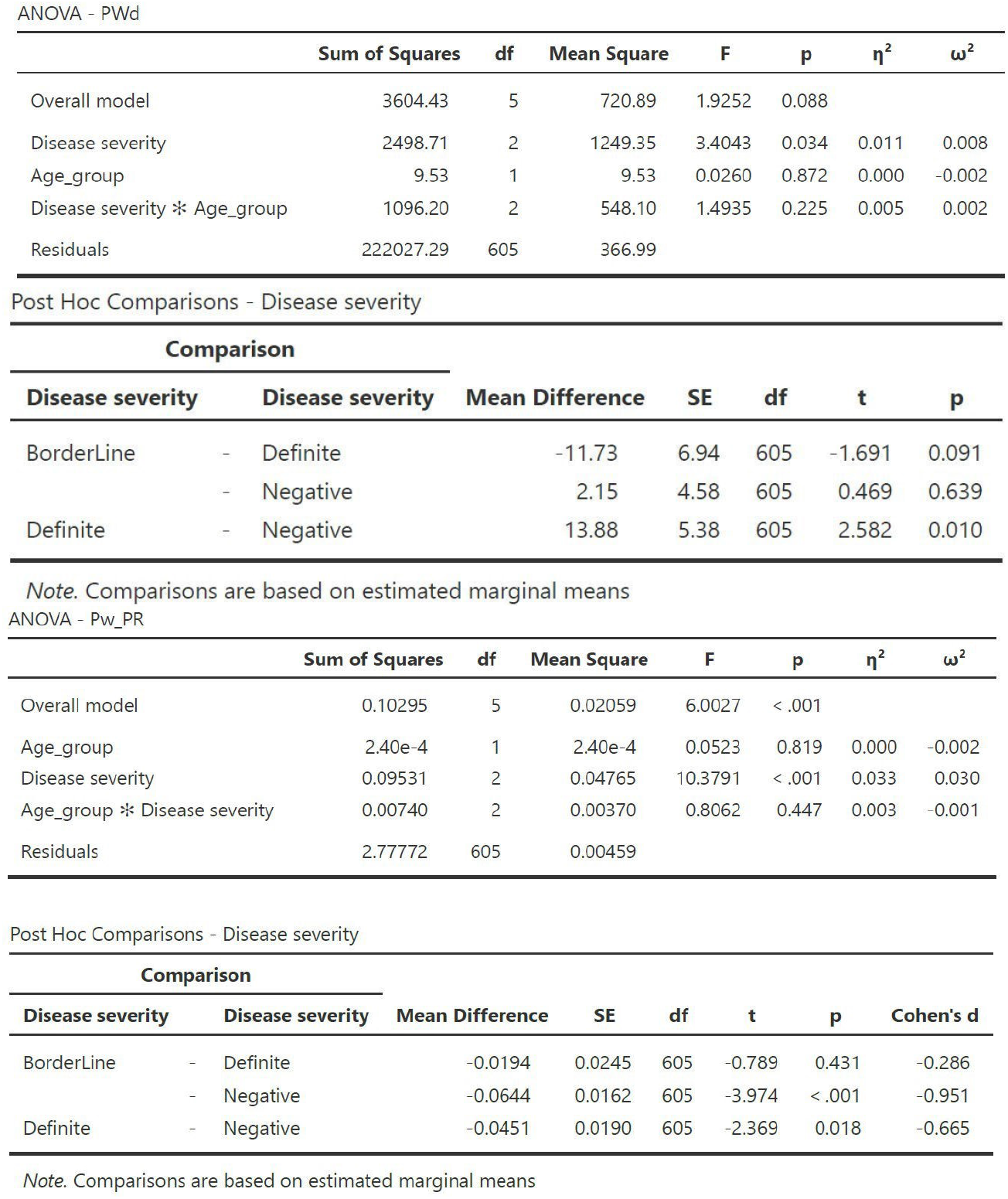

